# Short Communication: Vitamin D and COVID-19 infection and mortality in UK Biobank

**DOI:** 10.1101/2020.06.26.20140921

**Authors:** Claire E Hastie, Jill P Pell, Naveed Sattar

**Author notes:** Joint senior authors (Jill P Pell, Naveed Sattar). **Address for correspondence** Dr Claire Hastie, Institute of Health and Wellbeing, University of Glasgow, 1 Lilybank Gardens, Glasgow, G12 8RZ.

## Abstract

**Purpose:** Vitamin D has been proposed as a potential causal factor in COVID-19 risk. We aimed to establish whether blood 25-hydroxyvitamin D (25(OH)D) concentration was associated with COVID-19 mortality, and inpatient confirmed COVID-19 infection, in UK Biobank participants.

**Methods:** UK Biobank recruited 502,624 participants aged 37-73 years between 2006 and 2010. Baseline exposure data, including 25(OH)D concentration, were linked to COVID-19 mortality. Univariable and multivariable Cox proportional hazards regression analyses were performed for the association between 25(OH)D and COVID-19 death, and poisson regression analyses for the association between 25(OH)D and severe COVID-19 infection.

**Results:** Complete data were available for 341,484 UK Biobank participants, of which 656 had inpatient confirmed COVID-19 infection and 203 died of COVID-19 infection. Vitamin D was associated with severe COVID-19 infection and mortality univariably (mortality HR=0.99; 95% CI 0.98-0.998; *p*=0.016), but not after adjustment for confounders (mortality HR=0.998; 95% CI=0.99-1.01; *p*=0.696).

**Conclusions:** Our findings do not support a potential link between vitamin D concentrations and risk of severe COVID-19 infection and mortality. Recommendations for vitamin D supplementation to lessen COVID-19 risks may provide false reassurance.

---

In the hunt for modifiable COVID-19 risk factors, vitamin D has gained a lot of attention both in the media and within the scientific community.^1^ Proponents of such a link cite a few early studies that present circumstantial evidence. They are either ecological, at an individual level but unable to fully adjust for potential confounders, or they measured vitamin D once patients were already hospitalised with COVID-19 which introduces reverse causation, as vitamin D is a negative acute phase reactant.^2^

Despite the sparse evidence on vitamin D in COVID-19,^3^ the UK government is now leading an urgent review into whether there is any link,^4^ and the Welsh COVID-19 risk assessment tool already includes vitamin D supplementation as part of its recommendations.^5^

We previously observed no evidence of an association between blood 25-hydroxyvitamin D (25(OH)D) concentration and testing positive for SARS-CoV-2 in hospital in UK Biobank once potential confounders were adjusted for.^6^ Importantly, some of the variables that were associated with increased COVID-19 risk in our sample, for example lower socioeconomic status, being Black or South Asian, or being overweight or obese, are also associated with lower vitamin D. This suggests that the positive findings of other studies may in part be due to inadequate adjustment. Another recent study of UK Biobank data replicated this finding,^7^ but it would be more informative to relate 25(OH)D concentration to COVID-19-related mortality.

In the current analysis, we therefore linked baseline 25(OH)D concentration in 341,484 UK Biobank participants with complete data on covariates to Death Register data. In the sample, 203 participants died due to COVID-19 infection. We explored whether vitamin D as a continuous measurement, or vitamin D deficiency (defined as <25 nmol/L), or vitamin D insufficiency (defined as <50 nmol/L), were associated with risk of COVID-19 death using Cox proportional hazards regression analysis. Multivariate models were adjusted for all measured confounders as detailed in the table legend.

The results followed the same pattern that we observed for COVID-19 infection (Table).^6^ Lower vitamin D concentration and vitamin D deficiency were both associated with higher risk of COVID-19 death univariably, but not after adjustment for potential confounders. Vitamin D insufficiency was not associated with risk of COVID-19 death univariably or multivariably. Furthermore, we repeated our previous analysis of the association between vitamin D and confirmed COVID-19 infection, using univariable and multivariable poisson regression of inpatient diagnosed infection. There were 656 confirmed inpatient COVID-19 cases. Again, vitamin D concentration and vitamin D deficiency were associated with COVID-19 infection univariably but not multivariably (Table).

**Table:**
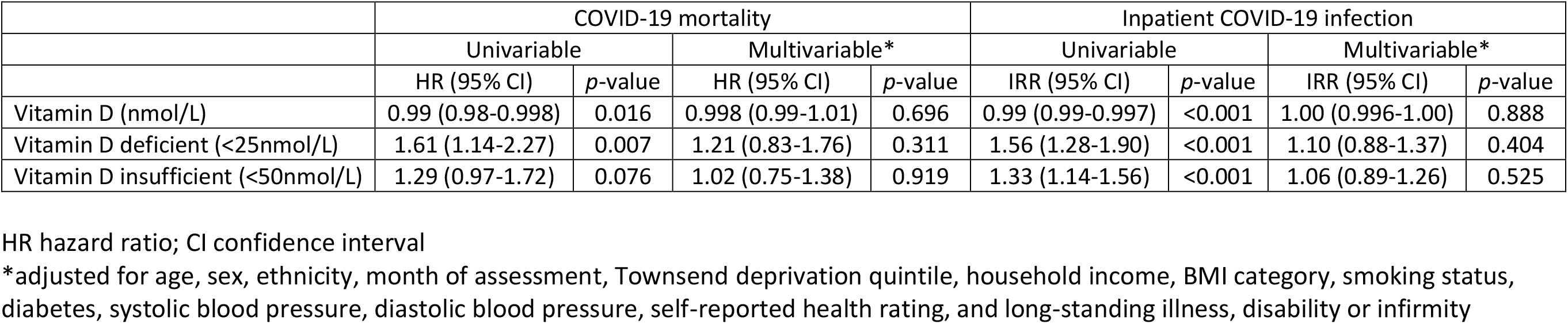
Association between vitamin D and confirmed COVID-19 mortality, and confirmed inpatient COVID-19 infection

|  | COVID-19 mortality |  |  |  | Inpatient COVID-19 infection |  |  |  |
| --- | --- | --- | --- | --- | --- | --- | --- | --- |
|  | Univariable |  | Multivariable* |  | Univariable |  | Multivariable* |  |
|  | HR (95% CI) | *p*-value | HR (95% CI) | *p*-value | IRR (95% CI) | *p*-value | IRR (95% CI) | *p*-value |
| Vitamin D (nmol/L) | 0.99 (0.98-0.998) | 0.016 | 0.998 (0.99-1.01) | 0.696 | 0.99 (0.99-0.997) | <0.001 | 1.00 (0.996-1.00) | 0.888 |
| Vitamin D deficient (<25nmol/L) | 1.61 (1.14-2.27) | 0.007 | 1.21 (0.83-1.76) | 0.311 | 1.56 (1.28-1.90) | <0.001 | 1.10 (0.88-1.37) | 0.404 |
| Vitamin D insufficient (<50nmol/L) | 1.29 (0.97-1.72) | 0.076 | 1.02 (0.75-1.38) | 0.919 | 1.33 (1.14-1.56) | <0.001 | 1.06 (0.89-1.26) | 0.525 |

HR hazard ratio; CI confidence interval
\*adjusted for age, sex, ethnicity, month of assessment, Townsend deprivation quintile, household income, BMI category, smoking status, diabetes, systolic blood pressure, diastolic blood pressure, self-reported health rating, and long-standing illness, disability or infirmity

The variables significantly associated with risk of COVID-19 mortality in multivariate analysis were age (HR=1.12; 95% CI 1.10-1.15; *p*<0.001 per year), male sex (HR=2.12; 95% CI 1.56-2.89; *p*<0.001), black ethnicity (HR=8.13; 95% CI 4.56-14.50; *p*<0.001), obesity (HR=1.68; 95% CI 1.11-2.56; *p*=0.015 compared with normal weight), socioeconomic deprivation (highest Townsend deprivation quintile compared with lowest HR=1.96; 95% CI 1.24-3.09; *p*=0.004), and diabetes (HR=1.96; 95% CI 1.34-2.86; *p*=0.001). These findings are consistent with other studies, lending strong external validity.

The main limitation of using UK Biobank for this analysis is the ∼10 year time period between baseline vitamin D measurement and COVID-19 infection. We examined concordance rates of vitamin D deficiency in a subsample of 15,473 participants who had measurements taken both at baseline and at a follow-up visit (on average 4.3 years later). Concordance in this group was 84%.

If there is a causal link, vitamin D supplements would present an appealingly cheap low risk intervention. However, currently there is no evidence that supplements will reduce risk of COVID-19 infection. NHS guidelines already recommend that all UK residents take vitamin D supplements in the winter, and furthermore that certain groups who are more likely to be deficient (for example those with darker skin) take them throughout the year.^8^ We await the results of randomised controlled trials to determine whether there should be any change to these guidelines and consequently clinical practice. For now, recommendations for vitamin D supplementation to lessen COVID-19 risks may cause little harm but could provide false reassurance.

## Data Availability

Data can be accessed via the procedures detailed in the UK Biobank website (https://www.ukbiobank.ac.uk/).

## Ethics approval

UK Biobank received ethical approval from the North West Multi-Centre Research Ethics Committee (REC reference: 16/NW/0274), and was conducted in accord with the principles of the Declaration of Helsinki.

